# Factors affecting Transition Success and Satisfaction from Child to Adult Mental Health Services in the UK

**DOI:** 10.1101/2022.11.29.22282875

**Authors:** Paris Williams, Kate Willoughby, Alison Bennetts, Valerie Brandt

**Author notes:** **Corresponding author:** Valerie Brandt, School of Psychology, Building 44, Highfield campus, SO17 1BJ Southampton. **Declaration of Interest:** None. **Ethics:** Written informed consent was obtained from all participants. This work complied with the ethical standards of the Helsinki Declaration and was approved by University of Southampton Ethics Board.

## Abstract

**Background:** Qualitative research has identified factors affecting transition from child mental health services (CAMHS) to adult mental health services (AMHS) but it is unclear which of these factors may lead to disengagement from the young persons’ view.

**Methods:** N= 272 participants (mean age = 20+/-2.31, 81% females) who had experience with the UK mental healthcare system (patients, carers, health workers) attempted the survey but only participants who had been treated in CAMHS were included in this study, resulting in a total N=144 (mean age = 19.8+/-2.3, 83% female). This study used a cross-sectional, quantitative survey assessing 12 pre-transition, 16 peri-transition, and 11 post-transition variables. The Client Attachment to Therapist Scale (CATS) was used to measure client attachment to CAMHS and AMHS therapists.

**Results:** Successful transition from CAMHS to AMHS was significantly predicted by using a helpful care plan, continuity of treatment between CAMHS and AMHS, and being engaged in a transition service. However, few clients were aware of transition services at the time of transition. Transition satisfaction was significantly predicted by the same variables. In addition, GP support during the transition, and a more secure attachment to AMHS therapists were associated with higher transition satisfaction.

**Conclusions:** The results suggest clients’ transition process might be significantly improved by focusing on useful individualised care plans, and ensuring continuity of treatment. Transition success and satisfaction could also be improved by making clients aware of and engaging them in transition services, involving GPs, and working on a secure attachment on the AMHS side.

## Introduction

In the UK, approximately 25,000 young people attempt the transition from child and adolescent mental health services (CAMHS) to adult mental health services (AMHS) each year (NHS, 2017). Transitions typically occur between age 16-18 years. At the same time, this adolescent period is often associated with increased psychological comorbidity (Murphy & P., 2012), and an increase in severity of pre-existing difficulties (Kessler et al., 2005). Ideally, any young person who required continued support for their mental health post 18 years, would experience a smooth transition to adult services. However, research suggests that out of the young people treated in child services, 61% stop receiving care once they turn 18 (3), and only 4% of those who transition to AMHS experience an ‘ideal’ transition (Singh et al., 2008). As a result, the transition process has been a central feature in government policy and initiatives. NICE guidelines (2016) highlighted the need for early planning, strength based and person-centred approaches, and ensuring joint working to enable better transition experiences (NICE, 2016). Furthermore, NHS England (2017) welcomed a new CQUIN (commissioning for quality and innovation), aiming to encourage improvements to the experiences and outcomes of young people transitioning (NHS, 2017).

There are various factors at the policy, organisation, service, and individual level that shape the complexities surrounding continuation of care. In a recent review, Hendrick (2020) outlined some of the challenges that CAMHS and AMHS face when ensuring policy is implemented into practise (Hendrickx et al., 2020). These complexities largely stem from cultural and systemic differences that exist between CAMHS and AMHS, including eligibility thresholds and waiting lists, which may increase the risk of young people being left without interim support (Hendrickx et al., 2020), lack of joint working, and poorly defined procedures (4-5). Yet, it is largely unclear, which specific factors could be prioritised to improve the transition experience for young people from the clients’ perspective.

With regard to service users’ views, Broad and colleagues (2017) conducted a qualitative thematic synthesis of eighteen qualitative studies that reflected the experiences of 253 young people (Broad et al., 2017). Several themes across the transition phase were highlighted as key for transition satisfaction and success. Some examples included specific qualities of the clinician (e.g. good listener, flexible, and supportive), the young person’s involvement in the planning process, and joint working between CAMHS and AMHS. However, it remains unclear, which of these themes are key in a successful and satisfactory transition. The present study used a cross-sectional quantitative survey, based on Broad and colleagues’ (2017) key themes to ascertain which factors differentiate between those service users that attempt to go on to AMHS but are unsuccessful, and those that attempt to transfer and are successful in receiving treatment in AMHS, based on Broad et al.’s research (Broad et al., 2017). The second aim of this study was to determine which key themes are related to young people’s satisfaction in their experience of moving from CAMHS to AMHS. Thirdly, the study explores how attachment to one’s therapist can influence the extent to which young people feel satisfied in their experience of transitioning between services.

## Methods

### Patient and Public Involvement

The study was developed based on a public engagement event for patients with Tourette syndrome (TicsConnect: https://www.southampton.ac.uk/news/2018/11/tourettes-tics-psychology.page). The event included a focus group with stakeholders (patients, carers, relatives) to develop study topics that mattered to patients. The research question was formulated during this focus group, while stakeholders were not involved in the study design. This was developed based on previous qualitative research. The study was also piloted to assess whether it was easy to understand and whether questions were unambiguous. Patients were able to comment on each part of the survey.

### Participants

In this study, n = 272 (n = 219 females) participants attempted the survey, consisting of two main sections: one related to transition satisfaction and one relating to attachment. To meet the inclusion criteria, participants were aged 16 or above, and had some experience with the British healthcare system (e.g. patient, past patient, relative, carer). Overall, N = 219 (84.6%) participants reported they had been in the healthcare system, n = 9 (3.5%) were parents or carers, and n = 31 (12%) said ‘other’ (family member, friend); N = 12 participants reported no relation to the mental health care system (‘student / female / adult’), and N = 2 participants reported they self-diagnosed and were therefore excluded.

N = 144 reported they had been in CAMHS (Table 1). Transition attempt was defined as people who reported they had wanted to transfer or tried to transfer from CAMHS to AMHS. N = 98 participants (N = 75 female, mean age = 19.8, SD = 2.63, range = 16-39; Table 2) had attempted transition. Transition success was defined as those people who reported they had attempted to transfer from CAMHS to AMHS and had been accepted into AMHS.

**Table 1.** Information about number of service users

|  | Was in<br>CAMHS | Diagnosed<br>in CAMHS | Currently treated<br>in AMHS | Currently assessed<br>in AMHS | Was aware of<br>transition<br>services |
| --- | --- | --- | --- | --- | --- |
| N (%) | 144 (57) | 83 (33) | 40 (16) | 31 (12) | 8 (5) |
|  | Wanted to<br>transfer | Tried to<br>transfer | Transitioned from<br>CAMHS to AMHS | Private healthcare | Transition<br>satisfaction<br>M (SD) |
| N (%) | 71 (28) | 46 (18) | 59 (23) | 78 (31%) | 1.60 (1.03)<br>Range: 0-4 |

Participants were asked questions with regard to their treatment in child mental health services (CAMHS) and adult mental health services (AMHS) and whether they had wanted and tried to transition, and whether the transition had been successful.

**Table 2.**
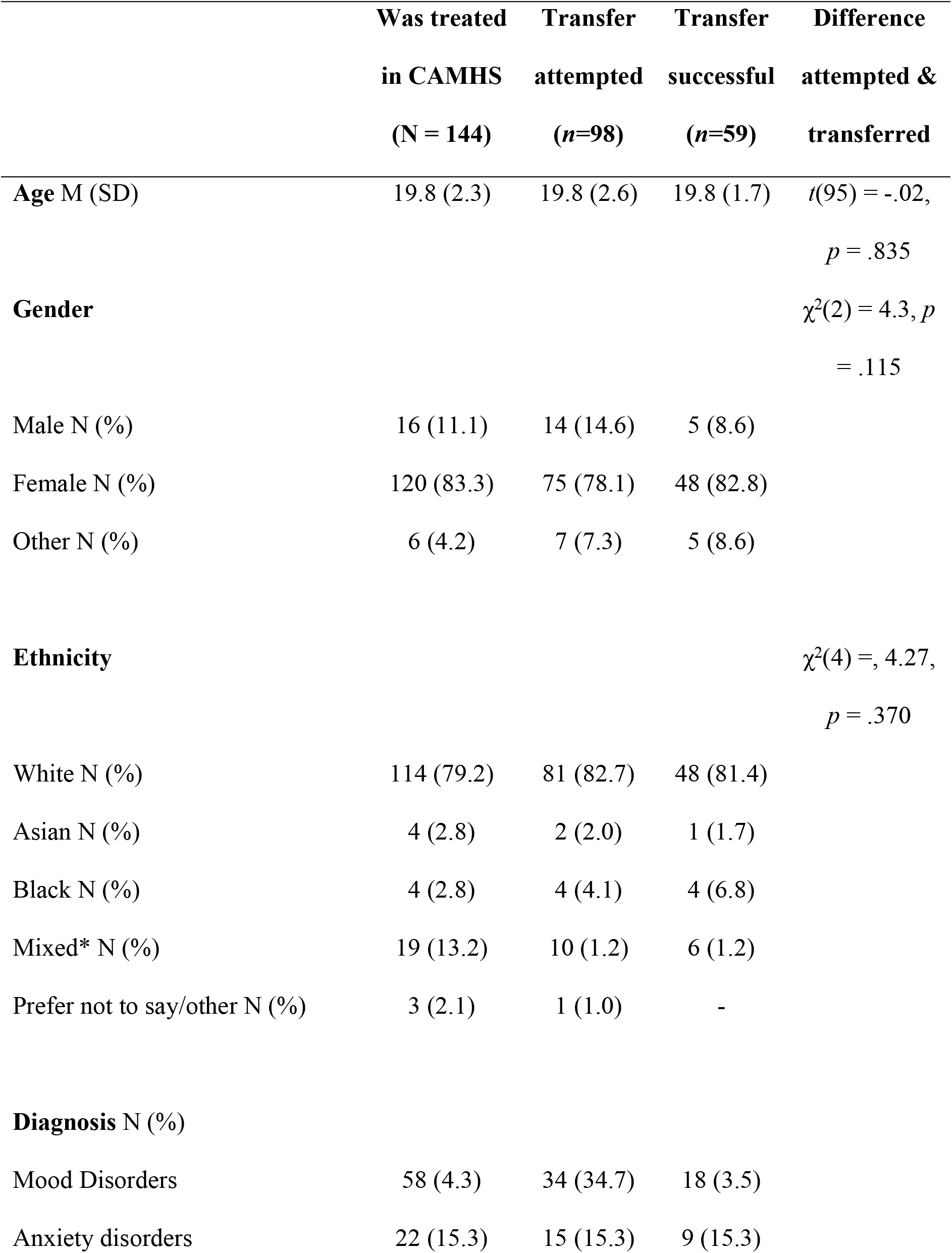
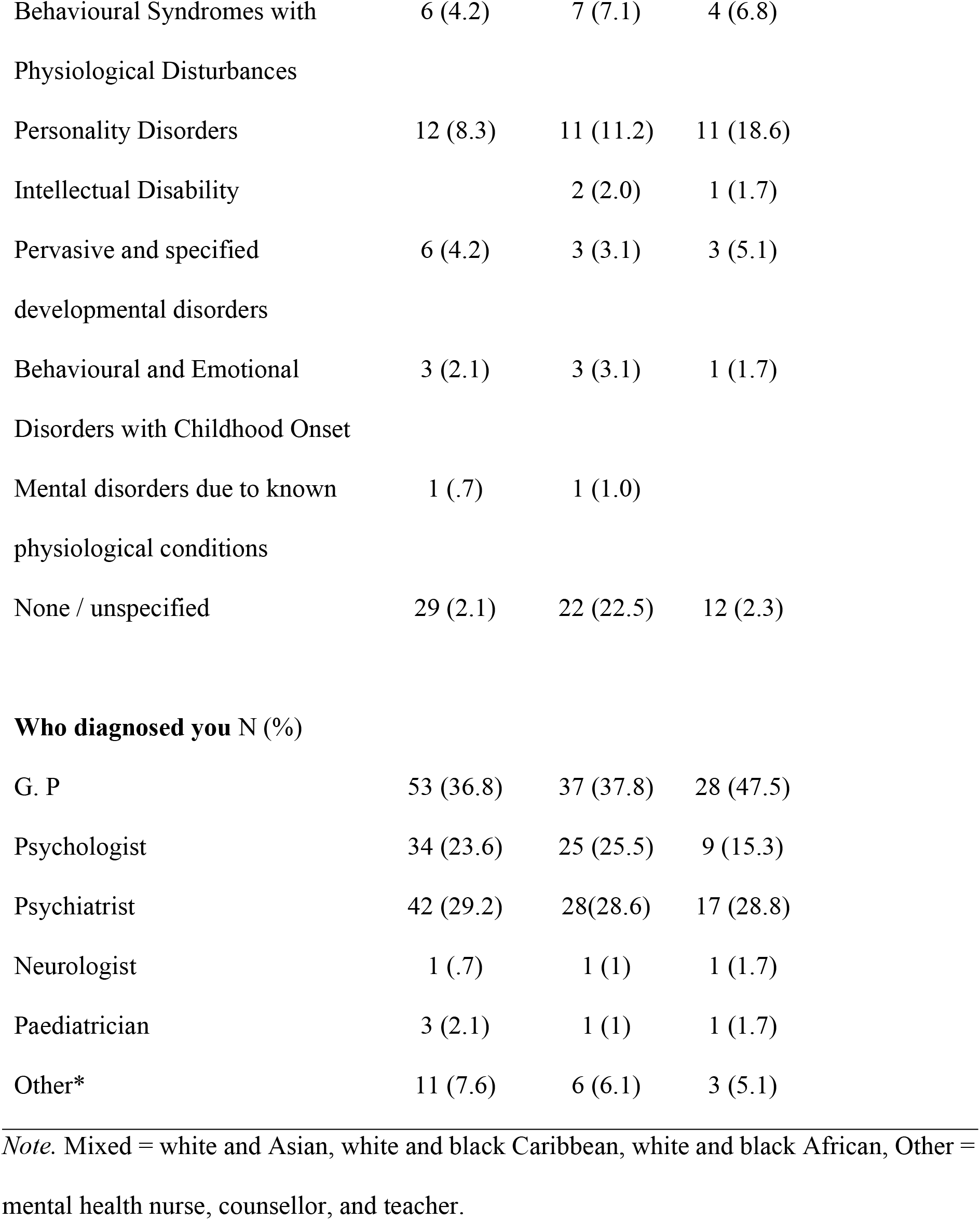
Demographic Characteristics of Participants

|  | **Was treated**<br>**in CAMHS**<br>**(N = 144)** | **Transfer**<br>**attempted**<br>**(n=98)** | **Transfer**<br>**successful**<br>**(n=59)** | **Difference**<br>**attempted &**<br>**transferred** |
| --- | --- | --- | --- | --- |
| **Age M (SD)** | 19.8 (2.3) | 19.8 (2.6) | 19.8 (1.7) | $t(95) = -.02,$<br>$p = .835$ |
| **Gender** | | | | $\chi^2(2) = 4.3, p$<br>$= .115$ |
| Male N (%) | 16 (11.1) | 14 (14.6) | 5 (8.6) |  |
| Female N (%) | 120 (83.3) | 75 (78.1) | 48 (82.8) |  |
| Other N (%) | 6 (4.2) | 7 (7.3) | 5 (8.6) |  |
| **Ethnicity** | | | | $\chi^2(4) = 4.27,$<br>$p = .370$ |
| White N (%) | 114 (79.2) | 81 (82.7) | 48 (81.4) |  |
| Asian N (%) | 4 (2.8) | 2 (2.0) | 1 (1.7) |  |
| Black N (%) | 4 (2.8) | 4 (4.1) | 4 (6.8) |  |
| Mixed* N (%) | 19 (13.2) | 10 (1.2) | 6 (1.2) |  |
| Prefer not to say/other N (%) | 3 (2.1) | 1 (1.0) | - |  |
| **Diagnosis N (%)** |  |  |  |  |
| Mood Disorders | 58 (4.3) | 34 (34.7) | 18 (3.5) |  |
| Anxiety disorders | 22 (15.3) | 15 (15.3) | 9 (15.3) |  |

|  |  |  |  |
| --- | --- | --- | --- |
| Behavioural Syndromes with<br>Physiological Disturbances | 6 (4.2) | 7 (7.1) | 4 (6.8) |
| Personality Disorders | 12 (8.3) | 11 (11.2) | 11 (18.6) |
| Intellectual Disability |  | 2 (2.0) | 1 (1.7) |
| Pervasive and specified<br>developmental disorders | 6 (4.2) | 3 (3.1) | 3 (5.1) |
| Behavioural and Emotional<br>Disorders with Childhood Onset | 3 (2.1) | 3 (3.1) | 1 (1.7) |
| Mental disorders due to known<br>physiological conditions | 1 (.7) | 1 (1.0) |  |
| None / unspecified | 29 (2.1) | 22 (22.5) | 12 (2.3) |

**Who diagnosed you N (%)**
|  |  |  |  |
| --- | --- | --- | --- |
| G. P | 53 (36.8) | 37 (37.8) | 28 (47.5) |
| Psychologist | 34 (23.6) | 25 (25.5) | 9 (15.3) |
| Psychiatrist | 42 (29.2) | 28(28.6) | 17 (28.8) |
| Neurologist | 1 (.7) | 1 (1) | 1 (1.7) |
| Paediatrician | 3 (2.1) | 1 (1) | 1 (1.7) |
| Other* | 11 (7.6) | 6 (6.1) | 3 (5.1) |

*Note.* Mixed = white and Asian, white and black Caribbean, white and black African, Other = mental health nurse, counsellor, and teacher.

Written informed consent was obtained from all participants. This work complied with the ethical standards of the Helsinki Declaration (World Medical Association, 2013) and was approved by University of Southampton Ethics Board.

Participants were recruited via volunteer sampling. Adverts were displayed on social media sites, the University of Southampton (UK) efolio system and through charities, inviting participants to complete the survey in exchange for study credit and a chance of winning one of ten £40 amazon vouchers. Participants were asked at the start if they had experience with the British mental health care system. If they clicked ‘no’, data collection was ended. Data was collected from April 2018 – January 2021.

### Measures

A Service Transition Survey was developed based on Broad and colleagues (2017) synthesis of qualitative research. They identified 13 themes that played a role in how patients viewed their transition from CAMHS to AMHS (Supplementary Table 1). They identified 3 pre-transition themes (themes that influenced clients’ transition experience before the transition process started), 5 peri-transition themes (themes that influenced clients’ transition experience during the transition), and 5 post-transition themes (themes that influenced clients’ transition experience after they had transitioned). Following standard scale construction guidelines (6), these themes were translated into items (Supplementary Table 1) that measured the agreement with each theme from 0 (strongly disagree) to 4 (strongly agree). The internal consistency for the pre-transition subscale (N_items_= 12, α =.91) and peri-transition subscale (N_items_= 16, α = .95) was excellent, and the internal consistency for the post-transition subscale (N_items_= 11, α = .82) was good.

A factor analysis with Varimax rotation on the pre-transition items revealed 2 factors: 1) ‘Quality of CAMHS clinician support (e.g. having a reliable, flexible and emotionally supportive clinician), and 2) ‘transition planning’ (i.e. perceived level of involvement in and preparation for the process; see Supplementary Table 2). Regarding the peri-transition items, factor analysis revealed 3 factors: 1) ‘involvement in transition’ (e.g. young person’s opinions and concerns considered, gradual and flexible transition, and regular contact with CAMHS and AMHS clinicians), 2) ‘transition structure’ (e.g. individualised care plans used and helpful, transition service engaged, continuity of treatment between CAMHS and AMHS), and 3) ‘G.P. support’ (practical and emotional support from GP during the transition; see Supplementary Table 3). The factor analysis on the post-transition items revealed 3 factors: 1) ‘positive AMHS experience’ (e.g. feeling supported by AMHS staff, autonomy regarding treatment decisions, information effectively transferred from CAMHS to AMHS), 2) ‘positive parent involvement’ (perceived level of choice regarding parental involvement, being comfortable with reduced parent involvement), 3) ‘negative AMHS experience’ (only one item: ‘I felt I had to keep repeating myself when I met with AMHS clinicians after my transition from CAMHS to AMHS’; see Supplementary Table 4).

Transition satisfaction was assessed using 5 items assessing satisfaction and happiness, as well as mental wellbeing during the transition, presented on a 5-point Likert scale (0-4) and factor analysis showed 1 factor (Supplementary Table 5). Reliability was good (N_items_ = 3, α =.85). The Client Attachment to Therapist Scale (CATS) was used to measure client attachment to their therapist. (8). This is a 36-item measure, asking clients to respond using a 6point scale ranging from strongly agree to strongly disagree. The measure is intended to assess client’s perceptions of the client-therapist relationship within an attachment theory framework. The CATS consist of three subscales including pre-occupied (I would like my therapist to feel closer to me), avoidant (It’s hard for me to trust my therapist) and secure attachment (my therapist is dependable). In our sample, the scale demonstrated good psychometric properties with internal consistency alphas of α = .88 for the CAMHS secure, α = .93 for the CAMHS avoidant, and α = .92 for the CAMHS preoccupied subscales. Similar alphas were found for the AMHS subscales of α = .77 for the secure, α = .90 for the avoidant, and .91 for the preoccupied subscales.

### Data Sharing Statement

The data is fully available from https://eprints.soton.ac.uk/451424/.

### Statistical Analyses

Several factor analyses with Varimax rotation were conducted to reduce the number of outcome variables (transition satisfaction items), and the number of predictors. Means were calculated across all items in each factor to build a limited number of predictors.

A binary logistic regression was used to assess the association between those who successfully transferred and those that did not (participants who wanted or tried to transfer but did not). Gender was included as a categorical predictor.

Linear regression analyses were used to investigate which of these factors significantly predicted transition satisfaction and how attachment style was related to transition satisfaction. Gender was included as a binary predictor.

Ns for each variable can be found in Supplementary Table 6. Missing values were not replaced because it cannot be assumed that all participants had experiences in all assessed variables, e.g. participants who did not transfer would not be able to comment on post-transfer items. Due to the same reason, missing values were excluded pairwise in the linear regressions.

## Results

The binary logistic regression model using pre-transition, and peri-transition factors to predict transition success was not quite significant (χ^2^(7) = 9.23, *p* = .24; Nagelkerke R^2^ =.17), indicating that other variable play an important role in the transition that were not considered in this model. A good peri-transition structure (using a helpful individualised care plan based on young people’s own needs, engaging in the transition process, and treatment continuity from CAMHS to AMHS) made transition success three times more likely (Table 3). A linear regression predicting transition satisfaction from all transition factors was overall significant (*F*(9,48) = 12.63, *p* < .001, R^2^ = .70). Involvement during transition and positive parent involvement in AMHS were significantly associated with transition satisfaction (Table 4). A binary logistic regression model predicting transition success from attachment to CAMHS or AMHS therapists showed no significant results (χ^2^(8) = 9.35, *p* = .31; Nagelkerke R^2^ =.29). A linear regression model predicting transition satisfaction from client attachment to therapist factors was overall significant (*F*(7,44) = 5.88, *p* < .001, R^2^ = .48); having a more secure relationship with one’s AMHS therapist was related to higher transition satisfaction (Table 5).

**Table 3.** Pre and peri-transition factors predicting transition success

|  | **B** | **S.E.** | **Wald** | ***p*** | **OR** | **CI lower** | **CI upper** |
| --- | --- | --- | --- | --- | --- | --- | --- |
| Constant | .03 | .81 | .00 | .975 | .98 |  |  |
| **Pre-transition factors** |  |  |  |  |  |  |  |
| Quality of CAMHS<br>clinician support | .06 | .30 | .04 | .843 | 1.06 | .58 | 1.93 |
| Transition planning | .03 | .43 | .00 | .951 | 1.03 | .45 | 2.37 |
| **Peri-transition factors** |  |  |  |  |  |  |  |
| Involvement in transition | -.58 | .59 | .97 | .324 | .56 | .18 | 1.77 |
| Transition structure | 1.14 | .48 | 5.56 | **.018** | 3.12 | 1.21 | 8.03 |
| GP support | .06 | .27 | .06 | .812 | 1.07 | .63 | 1.82 |
| **Gender** |  |  |  |  |  |  |  |
| Female |  |  | 3.29 | .193 |  |  |  |
| Male | .92 | 1.22 | .57 | .449 | 2.51 | .23 | 27.19 |
| Other gender | -1.60 | 1.00 | 2.55 | .110 | .20 | .03 | 1.44 |

The binary logistic regression model using pre-transition, and peri-transition factors to predict transition success explained 17% of the variation in outcome. Significant effects are highlighted in bold. N = 72.

**Table 4.** Regression models predicting transition satisfaction

|  | **B** | **SE** | **$\beta$** | ***t*** | ***p*** | **CI**<br>**lower** | **CI upper** |
| --- | --- | --- | --- | --- | --- | --- | --- |
| (constant) | -.08 | .56 |  | -.13 | .894 | -1.21 | 1.05 |
| **Pre-transition factors** |  |  |  |  |  |  |  |
| Quality of CAMHS clinician support | .05 | .09 | .05 | .55 | .585 | -.13 | .23 |
| Transition planning | .10 | .15 | .10 | .69 | .495 | -.19 | .39 |
| **Peri-transition factors** |  |  |  |  |  |  |  |
| Involvement in transition | .11 | .18 | .10 | .63 | .529 | -.24 | .46 |
| Transition structure | .33 | .14 | .33 | 2.41 | **.020** | .06 | .61 |
| GP support | .18 | .08 | .24 | 2.26 | **.028** | .02 | .35 |
| **Post-transition factors** |  |  |  |  |  |  |  |
| Positive AMHS experience | .16 | .10 | .18 | 1.63 | .109 | -.04 | .37 |
| Positive Parent Involvement | .01 | .09 | .01 | .07 | .942 | -.18 | .19 |
| Negative AMHS experience | -.14 | .08 | -.17 | -1.82 | .075 | -.29 | .02 |
| **Gender** |  |  |  |  |  |  |  |
| Male | .27 | .23 | .10 | 1.16 | .250 | -.20 | .74 |

**Table 5.** Regression models predicting transition satisfaction from attachment to therapist

|  | **B** | **S.E.** | **$\beta$** | ***t*** | ***p*** | **CI lower** | **CI upper** |
| --- | --- | --- | --- | --- | --- | --- | --- |
| (constant) | -1.55 | 1.01 |  | -1.54 | .130 | -3.58 | .48 |
| **CAMHS Clinician** |  |  |  |  |  |  |  |
| Secure attachment to therapist | .24 | .17 | .28 | 1.42 | .164 | -.10 | .59 |
| Avoidant attachment to therapist | .08 | .16 | .10 | .51 | .614 | -.25 | .41 |
| Preoccupied attachment to therapist | .07 | .13 | .10 | .58 | .565 | -.18 | .33 |
| **AMHS Clinician** |  |  |  |  |  |  |  |
| Secure attachment to therapist | .39 | .14 | .48 | 2.80 | **.008** | .11 | .68 |
| Avoidant attachment to therapist | -.03 | .15 | -.04 | -.22 | .827 | -.33 | .27 |
| Preoccupied attachment to therapist | .20 | .14 | .24 | 1.42 | .163 | -.08 | .47 |
| Gender | .30 | .33 | .11 | .93 | .357 | -.35 | .96 |

Linear regression model with transition satisfaction as the dependent variable. Significant effects are highlighted in bold.

## Discussion

Having an individualised care plan that was utilised and helpful during the transition, being engaged in a transition service, and continuity of treatment between CAMHS and AMHS made young people more satisfied and made their transition three times more likely to be successful. In addition, people who felt more emotionally and practically supported by their GP were more satisfied with their transition. It should also be noted that the overall model with regard to transition success was not significant, indicating that the themes identified in previous literature as important from the clients’ perspective (Broad et al., 2017) did not contribute a lot to understanding what made a transition successful but rather what determines transition satisfaction. The results also show that the type of attachment to CAMHS clinicians did not play a role for transition success but that a less secure attachment to AMHS clinicians was associated with lower satisfaction with the transition process.

A strength of this study is that it considers service users’ views in a quantitative way and is therefore able to pinpoint key factors that are important in a good and successful transition from the client perspective. Limitations include that we used a convenience sample and this also led to more women taking part in the study than men, limiting its’ generalizability, although gender was controlled for in the analyses. One advantage of the convenience sample was that patients were not pre-selected by either of the studied services, making it unlikely that selection biases played a role. Those people who engaged with a transition service had a higher likelihood of a successful transition but it is unclear whether these transition services were standalone services (e.g. third sector voluntary) or embedded within existing CAMHS or AMHS.

Our results are in line with previous research from the TRACK study, showing that many young people transfer successfully from CAMHS to AMHS, without experiencing ‘good transitional care’ (Moli Paul et al., 2013). With regard to transition services, transition workers have been found to be key in enabling a smoother and successful transition process, leading to better engagement rates (Singh et al., 2010), supporting young people at both the emotional and practical level (Haywood-Bell et al., 2016). Similarly, treatment in AMHS aiming to continue what the young person had previously been working on in CAMHS was important from the service users’ point of view. Getting to know the service user and working in line with their best interests and treatment goals appears to be facilitated by joint working between CAMHS and AMHS (Singh et al., 2010). Since 2017, joined working between the services and with the service user is a point that is specifically encouraged by an NHS CQUIN (NHS, 2017). The CQUIN recommends a transition plan, including meetings involving the young person, a key worker from the sending service, and a ‘point of contact’ from the receiving service (NHS, 2017). This might enhance transition planning involvement of the young person, and continuity of treatment across services. Our results regarding attachment stand in contrast to previous qualitative studies, which have suggested that the relationship young people have with their clinician, may play a role in experiencing a better transition (Lindgren et al., 2013; Wheatley et al., 2013). While qualitative research takes into account individual viewpoints, the quantitative nature of this study allows us to draw conclusions that are more generalizable.

The importance of GP support for transition satisfaction may reflect the need for additional support, outside of community mental health teams to negotiate the move between CAMHS and AMHS (Lindgren et al., 2013). Interestingly, our results suggest that there is a lack of awareness about transition services in clients that could be addressed systematically during transfer planning to make a transition more likely to be successfully completed. Transitioning between services is often accompanied with a loss of familiar and safe relationships to the formation of new ones. Therefore, ensuring staff in AMHS are sensitive to the attachment needs of the young person is important and in line with providing person centred planning. Preparatory sessions could be one way to facilitate relational and emotional safety, with young people having focussed time to meet with their new clinician to start developing new attachments, whilst gradually ending attachments with their CAMHS clinician. Future research should investigate what makes an individual care plan helpful or unhelpful to clients. Merely having a care plan did not improve likelihood of transition success. It should also be determined which transition services are most helpful in ensuring a higher likelihood of successful transition.

## Data Availability

The data is fully available from https://eprints.soton.ac.uk/451424/.

## Acknowledgments

We would like to thank Abilgail Targett for helping with the development of the survey and for piloting the survey.

## Contributorships

VB, KW, AB and PW designed the study, PW conducted the study, ran one set of analyses, and wrote the first draft of the paper, VB ran an independent set of analyses, VB, KW, and AB critically revised the manuscript.

